# Perception of the learning environment and its relationship with psychological distress in a cohort of preclinical medical and dental students in Nigeria: A cross-sectional study

**DOI:** 10.1101/2025.10.30.25339141

**Authors:** Adeniyi Abraham Adesola, Abigail Olawumi Oyedokun, Moses Ojomakpenen Ojile, Moses Dabor, Emmanuel Oluyinka Idowu

**Author notes:** **AAA:**; **AOO:**; **MOO:**; **DM:**; **EOI:**.

## Abstract

**Background:** The learning environment strongly influences academic outcomes and wellbeing. This study assessed perception of the preclinical learning environment at the College of Medicine, University of Ibadan and explored associations with psychological distress.

**Methods:** A cross-sectional study was conducted among 187 second and third-year medical and dental students in a tertiary institution in south-western Nigeria using convenient sampling. An online structured self-administered questionnaire was adapted from the Dundee Ready Education Environment Measure (DREEM) inventory and the Kessler Psychological Distress Scale (K10). Data were analysed using descriptive statistics and non-parametric tests, and correlations were assessed with Spearman’s rho at a 95% confidence level.

**Results:** The overall mean DREEM score was 124.6 ± 23.7, reflecting a more positive than negative perception of the learning environment. The mean scores for the different domains were Perception of Learning 29.0 ± 7.7, Perception of Teachers 30.0 ± 5.9, Academic Self-Perception 20.7 ± 5.7, Perception of Atmosphere 29.1 ± 6.5 and Social Self-Perception 15.9 ± 4.0. The mean score on the K10 was 21.7, with 43.3% of respondents classified as likely well, 29.0% with mild distress, 13.4% with moderate distress and 13.9% with severe distress. Older students, male students and those in their third year reported higher DREEM scores, while higher academic grades were also associated with more positive perception. Psychological distress showed a negative correlation with the overall DREEM score (*r*L = - 0.162; p = 0.027) and with several of its domains, suggesting that higher levels of distress were linked to less favourable perception of the learning environment.

**Conclusion:** Preclinical students at the University of Ibadan overall perceive their learning environment more positively, but substantial levels of psychological distress exist and are associated with poorer perception. Institutions should prioritise student wellbeing and strengthen feedback to enhance both educational climate and mental health.

**Strengths and limitations of this study:**

- The study focuses on preclinical medical and dental students, a group often underrepresented in research on the learning environment.
- It is the first known study in Nigeria to explore the relationship between learning environment perceptions and psychological distress using validated tools.
- The use of two well-established instruments — the DREEM inventory and Kessler Psychological Distress Scale — enhances methodological validity and comparability with international research.
- The study design was cross-sectional and limited to a single institution, which may restrict generalisability to other medical schools.
- Data were self-reported and collected during a specific academic period, which may introduce response and contextual biases.

## Introduction

Academic learning is a dynamic and multifaceted process shaped by multiple interacting factors that collectively determine a learner’s journey. These factors are broadly categorised into internal and external components. Internal factors encompass innate abilities such as intelligence quotient (IQ), cognitive skills, and intrinsic motivation, all of which influence how effectively a student assimilates, understands, and retains knowledge (1,2). Personal efforts, including reading habits, consistency, resilience, and time management, also play critical roles in academic success (3,4).

Notably, external influences play an equally crucial role in shaping students’ educational experiences, with the learning environment being one of the most significant. The learning environment can be defined as the physical, psychological, academic, and social conditions under which education occurs. It directly impacts academic performance, well-being, motivation, and professional development (5,6). A supportive environment encourages critical thinking, resilience, and career satisfaction, while an unsupportive one may lead to disengagement, poor performance, and emotional distress (7).

Several interrelated factors define the learning environment in medical education. Access to well-equipped libraries, lecture theatres, laboratories, and technological resources is fundamental. Equally vital are effective curriculum delivery, engaging teaching methods, and clear course objectives that enhance understanding (8). In addition, the attitude and competence of lecturers, assessment design, feedback mechanisms, and institutional policies all influence how students experience their education (8,9). Understanding the perception is essential for improving medical education and fostering environment that promote academic excellence and student well-being.

Evidence from previous research reinforces the association between positive perception of the learning environment and improved outcomes. A 2024 study among university students in China, as well as one among medical students at Jimma University Medical Centre, Ethiopia, revealed that those with positive perception often achieve higher academic performance and demonstrate greater psychological resilience (3,10). The latter study also noted better learning environment perception among females compared to males, which is similar to findings among year-one clinical students (10). This interesting pattern may suggest that certain student groups maintain more optimistic outlooks or experience stronger support networks early in training. All in all, environment that promote respect, constructive feedback, and inclusivity enhance both academic success and psychological well-being. Additionally, socially connected and well-supported students tend to perform better academically and often report lower levels of psychological distress (11,12).

Across Africa, particularly in Nigeria, infrastructural and systemic challenges make the study of learning environment especially important. Olusanya et al. (2019) found that although dental residents in a Nigerian training institution were satisfied with certain aspects of their training, they raised concerns about poor feedback systems, unapproachable teachers, and inadequate infrastructure (13). Similarly, medical students in Ethiopia identified overcrowded lecture halls, insufficient laboratory facilities, and strained faculty–student relationships as barriers to learning (10,14). In Nigeria, however, factors such as off-campus accommodation, limited extracurricular opportunities, and heavy academic workloads often restrict social interaction and reduce the overall quality of the learning experience (15,16).

Within Nigerian medical and dental education, where regulatory standards are stringent and curricula are demanding, the value of a positive learning environment is particularly significant (17,18). Preclinical students, who are in the formative phase of professional development, are especially sensitive to the educational climate (19). At the University of Ibadan, preclinical students (in years 2 and 3) engage with a rigorous curriculum, the majority of which comprises Human Anatomy, Physiology, and Biochemistry. They often lack adequate coping mechanisms, which increases the risk of burnout and disengagement. Many students struggle to appreciate the relevance of basic science courses to future clinical practice, as they experience surface learning and reduced motivation early in their training (15,20). Students may have multiple individuals in a dissection group or laboratory practical, with limited access to visual aids that could enhance retention. The University of Ibadan is the pioneer medical school in Nigeria, and the perception of its students may reflect wider patterns across the country’s medical institutions (21).

Several validated tools have been developed to objectively evaluate students’ experiences and well-being in medical education. The Dundee Ready Educational Environment Measure (DREEM), developed by Roff and colleagues in 1997 through a Delphi process involving international medical educators, is one of the most widely used instruments for assessing the educational climate in health professional schools (22,23). It has been validated across diverse cultural and institutional contexts, consistently demonstrating strong internal consistency and reliability. A cross-sectional study conducted in Jos, Northern Nigeria, which assessed the perception of clinical medical students, revealed that their overall perception of the learning environment was more positive than negative, with social self-perception scoring lower than academic self-perception (24). Another study among private medical schools in South-Eastern Nigeria also reported more positive than negative perception, with the social domain having the lowest mean score (25).

Similarly, the Kessler Psychological Distress Scale (K10), developed by Kessler and Mroczek in the 1990s, has been extensively employed in national and international health surveys as a brief yet robust screening tool for anxiety and depressive symptoms (26). It was used in a 2022 study among Nigerian healthcare workers during the COVID-19 pandemic, which recorded high levels of psychological distress (27). Taken together, the DREEM and K10 provide complementary insights. While DREEM evaluates students’ perception of their educational environment, the K10 captures the emotional and psychological dimensions of their well-being. Their combined use therefore enables a more holistic understanding of how the learning environment influences the mental health and academic experiences of medical and dental students.

Despite the recognised importance of the learning environment, local data on perception among preclinical medical and dental students in Nigeria, and particularly at the University of Ibadan, remain limited. Most existing Nigerian studies focus on clinical students or postgraduate trainees, leaving the unique experiences of preclinical students underexplored (13,28,29). Studies that collect data after students advance into clinical training risk recall bias, as subsequent experiences may influence their recollections.

Capturing perception while students are still in the preclinical phase provides more immediate and accurate insights into the strengths and weaknesses of the educational system. This study is novel in that it explored the views of students actively engaged in preclinical training, rather than relying on retrospective accounts. It sought to assess the perception of the learning environment among preclinical medical and dental students at the University of Ibadan, identify factors that promote effective learning, and those that require improvement. It contributes to the creation of a more supportive, inclusive, and effective educational environment for future healthcare professionals, as it provides nuanced findings that can inform targeted educational reforms.

## Methods

### Study site and settings

This study was conducted among undergraduate preclinical students enrolled in the Bachelor of Medicine, Bachelor of Surgery (MBBS) and Bachelor of Dental Surgery (BDS) degree programmes at the College of Medicine, University of Ibadan, Nigeria. Both programmes span six years and are divided into three phases: preliminary, preclinical, and clinical. The preliminary and preclinical students are based on the main University of Ibadan campus, while the clinical students are located in the College of Medicine complex within the University College Hospital (UCH), Ibadan. The focus of this research was the preclinical phase, which comprises three semesters of basic medical sciences in Anatomy, Physiology, and Biochemistry.

### Study design, sample size and sampling technique

A descriptive cross-sectional design was adopted to assess students’ perception of their learning environment at a single point in time. This design was appropriate for addressing the study’s descriptive objectives, such as measuring the distribution of positive and negative perception, and for exploring associations between sociodemographic characteristics, academic indices, and psychological distress.

The target population included all second- and third-year medical and dental students currently enrolled in the preclinical phase of the MBBS and BDS programme of study. Based on institutional records, approximately 360 students (180 per level) were eligible. Using Cochran’s formula with finite population correction, a minimum sample size of 186 was estimated to provide sufficient statistical power. Data were collected using a convenience sampling technique, yielding a final sample of 187 respondents, which transcended the threshold sample size calculated.

### Data collection procedure, inclusion and exclusion criteria

Data was collected through a structured, self-administered online questionnaire developed using Google forms. The link was distributed electronically via the students’ official class WhatsApp platforms of the second- and third-year medical and dental students. The first page of the questionnaire included a brief introduction explaining the purpose of the study, ensuring confidentiality, and obtaining informed consent. Only students who gave consent proceeded with filling the questionnaires. Eligible participants included all second- and third-year medical and dental students who were fully registered in the College of Medicine and willing to provide informed consent. Students who had deferred admission, allied health students, and those who declined participation were excluded from the study.

### Survey instrument

The questionnaire consisted of three main sections:

1. Sociodemographic and academic data: This section collected key characteristics of the respondents, including age, sex, academic level, degree programme enrolled into, accommodation type, and average academic score range.
2. Perception of learning environment: This was assessed using the Dundee Ready Education Environment Measure (DREEM), a validated 50-item instrument developed by Roff et al. (1997) to evaluate the educational climate in health professional training programmes (30). Each item was rated on a five-point Likert scale, with scores of 4 for strongly agree, 3 for agree, 2 for neutral, 1 for disagree, and 0 for strongly disagree. The DREEM items are categorised into five domains reflecting different aspects of students’ perception: Students’ Perception of Learning (SPL), Students’ Perception of Teachers (SPT), Students’ Academic Self-Perception (SASP), Students’ Perception of Atmosphere (SPA), and Students’ Social Self-Perception (SSSP). In this study, the internal consistency of these domains was satisfactory, with Cronbach’s alpha coefficients of 0.863, 0.798, 0.865, 0.788, and 0.608, respectively, while the overall scale demonstrated excellent reliability with a Cronbach’s alpha of 0.903. The total possible DREEM score is 200, with higher scores indicating a more positive perception of the learning environment.
3. Psychological distress: This was screened using the Kessler Psychological Distress Scale (K10). The K10 is a 10-item tool developed by Professors Ron Kessler and team in 1992 to be used as a screening tool for non-specific distress (31). Items are scored on a five-point Likert scale ranging from 1 (none of the time), 2 (a little of the time), 3 (some of the time), 4 (most of the time), to 5 (all of the time). The K10 demonstrated good internal consistency in this study, with a Cronbach’s alpha coefficient of 0.895.

### Data analysis

Data obtained was cleaned via Microsoft Excel and subsequently imported to IBM SPSS version-27 for analysis. Descriptive statistics such as frequencies, percentages, means, and standard deviations were used to summarise sociodemographic and academic characteristics, as well as students’ responses to the individual items of the DREEM inventory and the Kessler Psychological Distress Scale. The DREEM items were scored on a five-point Likert scale, with negatively worded items reverse-coded prior to analysis - items 4, 8, 9, 17, 25, 35, 39, 48 and 50 were reverse scored. Domain scores for Students’ Perception of Learning (SPL), Students’ Perception of Teachers (SPT), Students’ Academic Self-Perception (SASP), Students’ Perception of Atmosphere (SPA), and Students’ Social Self-Perception (SSSP) were computed according to standard DREEM guidelines (Table 1). The total DREEM score, representing the overall perception of the learning environment, was obtained by summing all domain scores. Interpretation of domain and overall scores followed Roff et al.’s established criteria, with higher scores indicating a more positive perception.

**Table 1:**
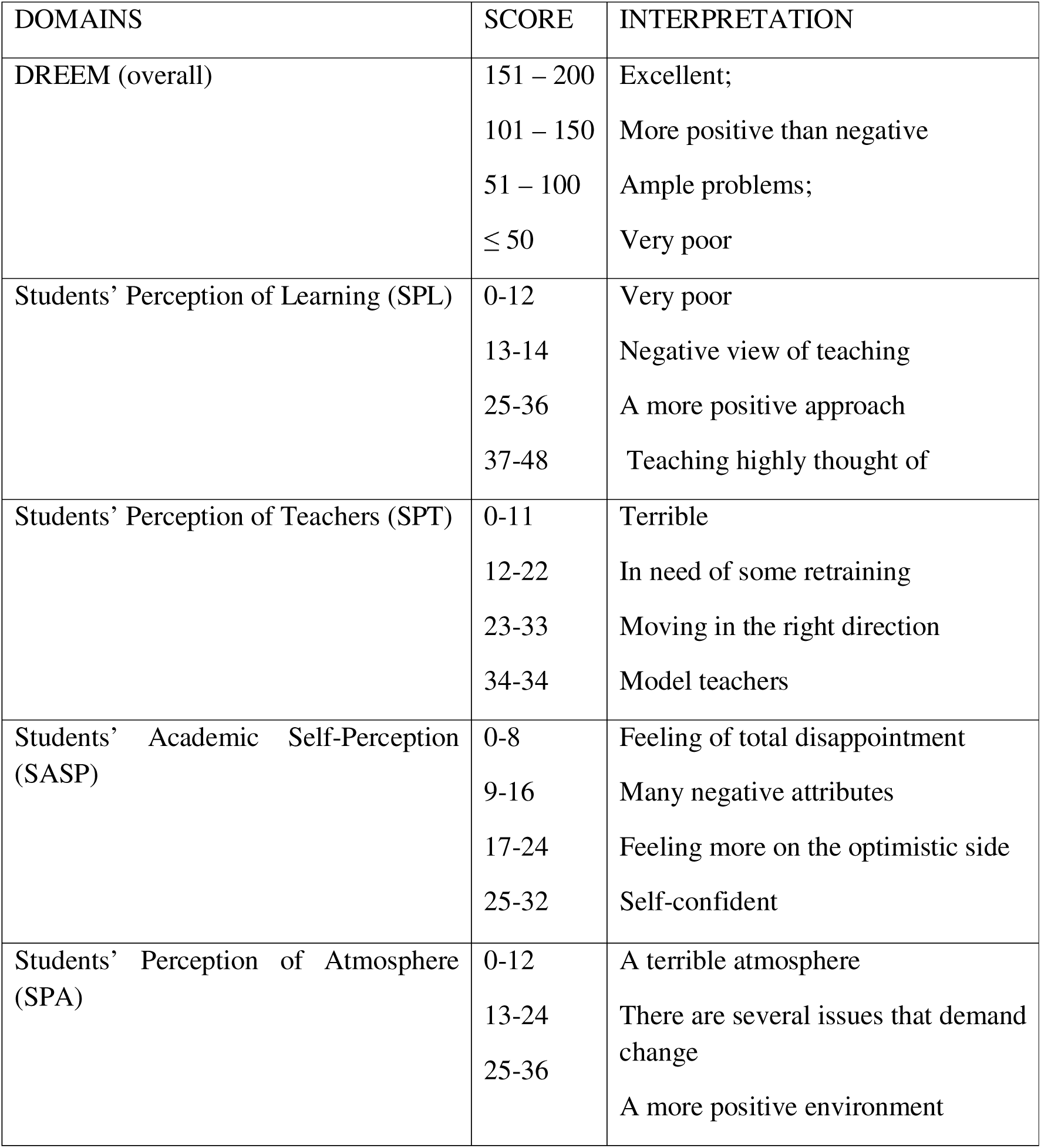
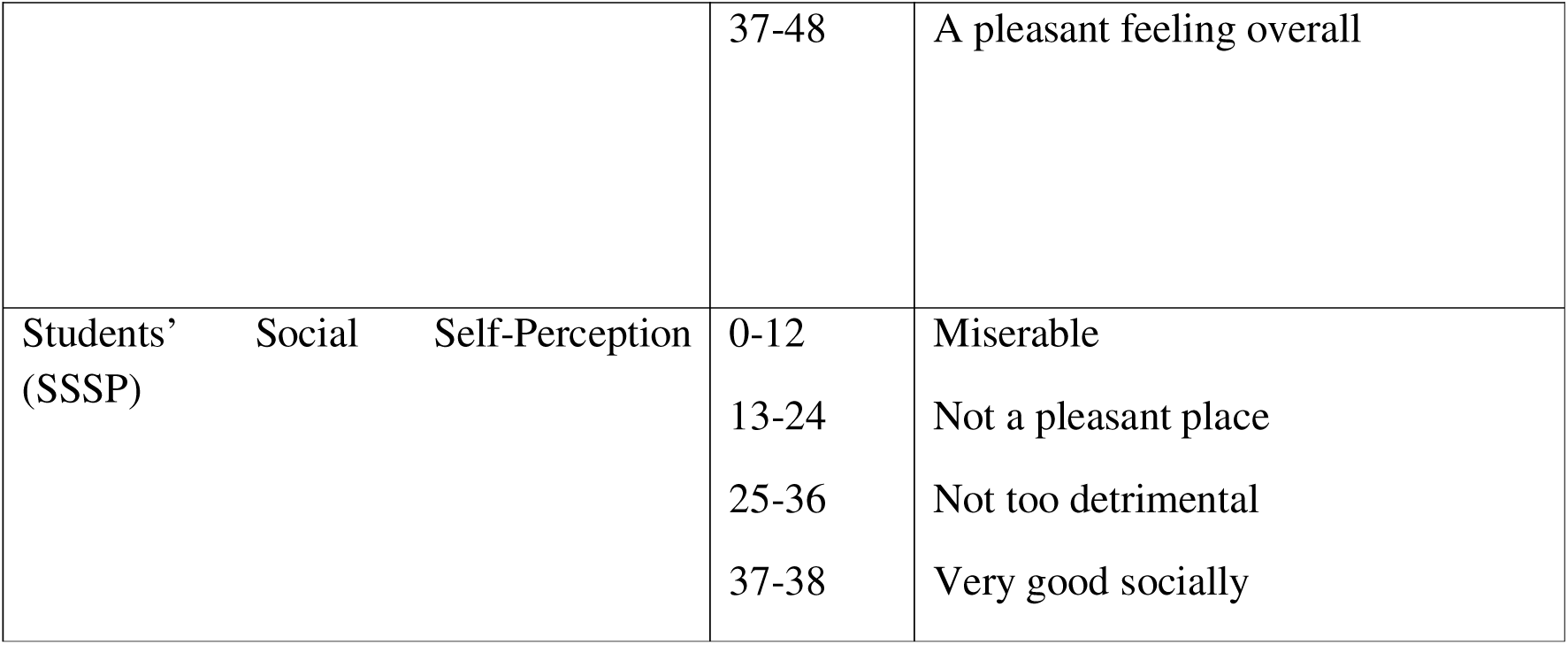
Guiding principle for determining the DREEM score and interpretation according to domain.

The Kessler Psychological Distress Scale (K10) was used to categorize respondents into different levels of psychological distress - likely well (scores 10 – 19), mild (scores 20 – 24), moderate (scores 25 – 29), and severe (scores 30 – 50). Mean Kessler scores were computed, and participants were grouped based on established cutoff points to determine the proportion of students experiencing mild, moderate, or severe distress. To examine associations between sociodemographic variables and DREEM domain scores, non-parametric tests were employed because the data did not meet the assumptions for normality. The Mann–Whitney U test was used for comparisons between two groups (e.g., gender, accommodation type, presence of mental illness), while the Kruskal–Wallis test was applied for comparisons involving more than two groups (e.g., age, academic grade, psychological distress categories). Where significant differences were found, post-hoc pairwise comparisons were performed to identify the source of variation. The relationship between psychological distress and DREEM domain scores was assessed using Spearman’s rank correlation coefficient. Statistical significance was set at p < 0.05 for all analyses. Results were presented using appropriate tables and summary statistics for clarity.

### Ethical considerations

Ethical approval for this study was obtained from the University of Ibadan/University College Hospital (UI/UCH) Ethics Committee. The study was conducted in accordance with the principles outlined in the Declaration of Helsinki.

## Results

### Sociodemographic and academic characteristics (Table 2)

The mean age of respondents was 21.75 ± 2.35 years, with most participants aged 22–26 years (51.3%). A greater proportion were male (70.6%), enrolled in Medicine and Surgery (84.5%), and predominantly resided on campus (81.8%). 5.9% of students reported being on treatment for a mental health condition, and 58.3% had an average academic grade of ≥70%. Regarding extracurricular engagement, 37.4% reported high engagement, while 33.2% and 29.4% had low and moderate engagement, respectively. On the Kessler Psychological Distress Scale, 43.3% were classified as *well*, 29 % as mildly distressed, 13.4% as moderately distressed and 13.9% as *severely distressed*. The mean Kessler score was 21.7, indicative of a moderate psychological distress score.

**Table 2:** Sociodemographic and academic characteristics of respondents.

| Variable | Category | Frequency | Percentage (%) |
| --- | --- | --- | --- |
| Age (years)<br>Mean (SD) = 21.75 (2.35) | 17 - 21 | 85 | 45.5 |
|  | 22 - 26 | 96 | 51.3 |
|  | 27 - 31 | 6 | 3.2 |
| Gender | Female | 55 | 29.4 |
|  | Male | 132 | 70.6 |
| Department | Medicine and Surgery | 158 | 84.5 |
|  | Dentistry | 29 | 15.5 |
| Academic level | 2nd year | 93 | 49.7 |
|  | 3rd year | 94 | 50.3 |
| Accommodation type | Off campus | 34 | 18.2 |
|  | On campus | 153 | 81.8 |
| Average academic grade | <50% | 2 | 1.1 |
|  | 50-59 | 28 | 15.0 |
|  | 60-69 | 48 | 25.7 |
| | $\geq 70$ | 109 | 58.3 |
| Mental illness | No | 176 | 94.1 |
|  | Yes | 11 | 5.9 |
| Extracurricular engagement | low | 62 | 33.2 |
|  | moderate | 55 | 29.4 |
|  | High | 70 | 37.4 |
| Kessler Distress Category<br>Mean (SD) = 21.7 (6.97). | Low distress | 81 | 43.3 |
|  | moderate distress | 55 | 29.4 |
|  | high distress | 25 | 13.4 |
|  | very high distress | 26 | 13.9 |

### Students’ perception of environment

A summary of the responses to the DREEM items has been presented in Table 3. Notably, some items recorded mean scores below 2.00, indicating areas that may require attention. Students reported that the enjoyment of studying was outweighed by stress (1.39), reflecting a need for a better balance between academic demands and wellbeing. In addition, students found the course boring at times (1.81), suggesting that teaching methods and course delivery could be made more engaging to sustain attention and motivation.

**Table 3.**
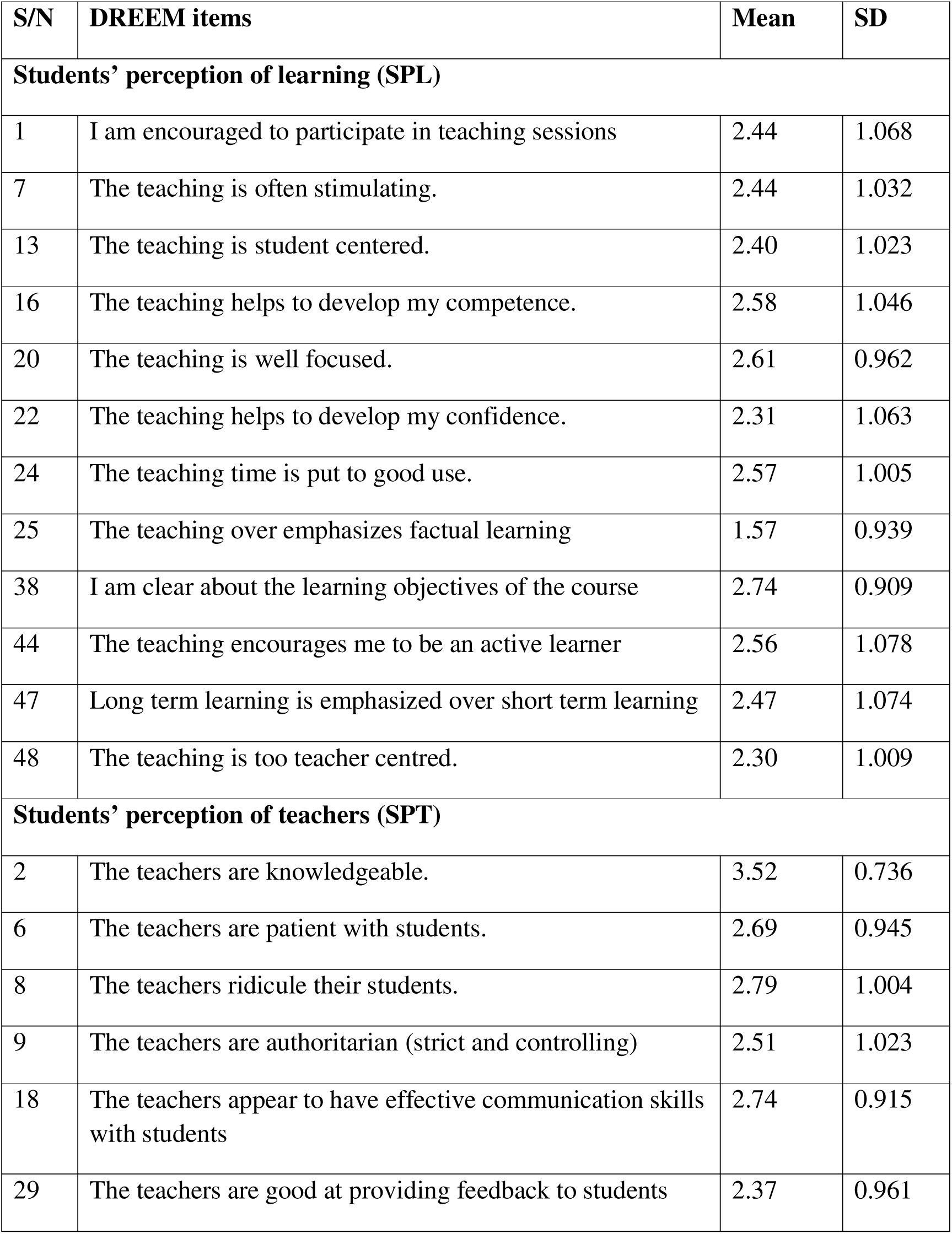
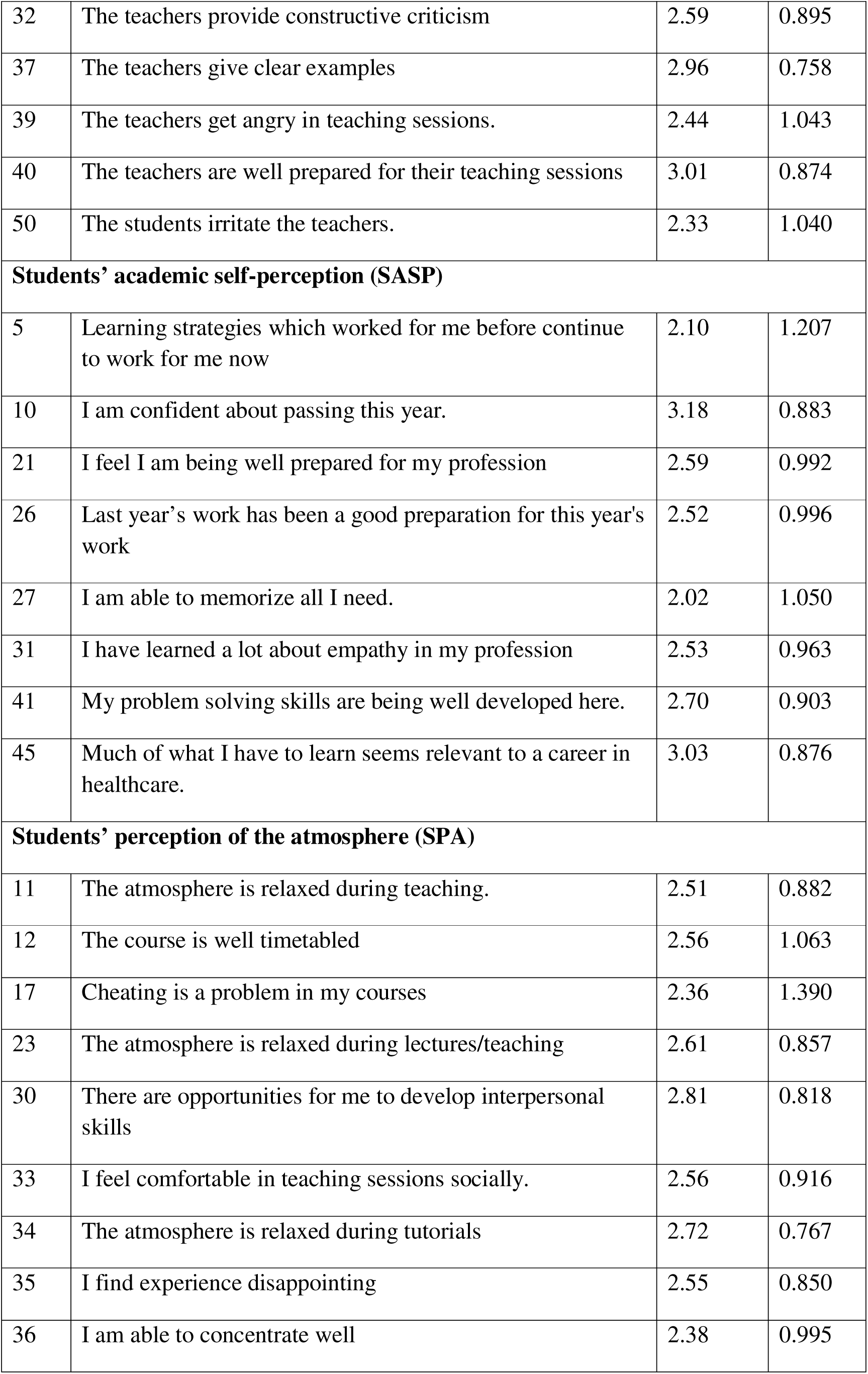
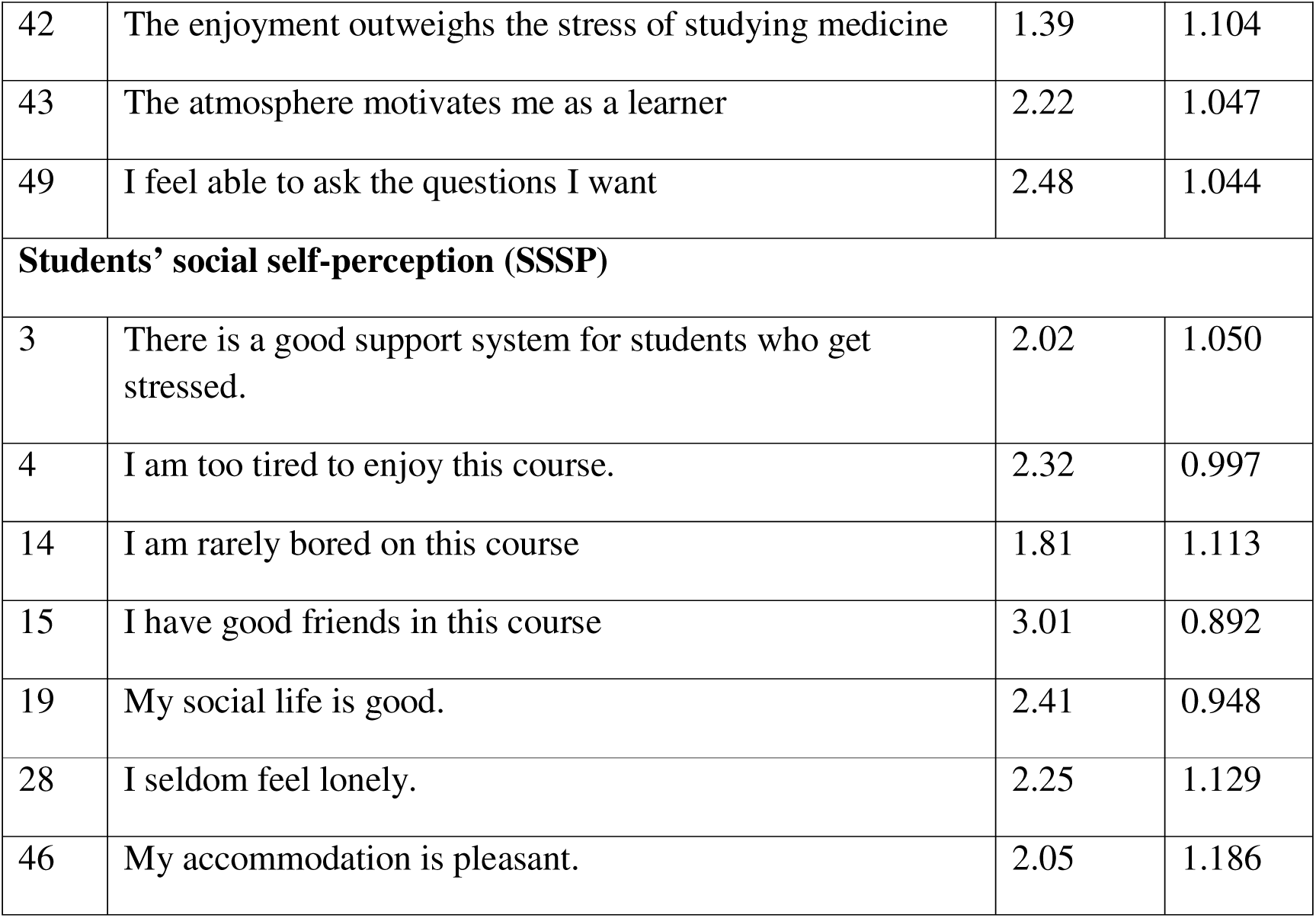
Characteristics of individual DREEM items.

| S/N | DREEM items | Mean | SD |
| --- | --- | --- | --- |
| **Students’ perception of learning (SPL)** |  |  |  |
| 1 | I am encouraged to participate in teaching sessions | 2.44 | 1.068 |
| 7 | The teaching is often stimulating. | 2.44 | 1.032 |
| 13 | The teaching is student centered. | 2.40 | 1.023 |
| 16 | The teaching helps to develop my competence. | 2.58 | 1.046 |
| 20 | The teaching is well focused. | 2.61 | 0.962 |
| 22 | The teaching helps to develop my confidence. | 2.31 | 1.063 |
| 24 | The teaching time is put to good use. | 2.57 | 1.005 |
| 25 | The teaching over emphasizes factual learning | 1.57 | 0.939 |
| 38 | I am clear about the learning objectives of the course | 2.74 | 0.909 |
| 44 | The teaching encourages me to be an active learner | 2.56 | 1.078 |
| 47 | Long term learning is emphasized over short term learning | 2.47 | 1.074 |
| 48 | The teaching is too teacher centred. | 2.30 | 1.009 |
| **Students’ perception of teachers (SPT)** |  |  |  |
| 2 | The teachers are knowledgeable. | 3.52 | 0.736 |
| 6 | The teachers are patient with students. | 2.69 | 0.945 |
| 8 | The teachers ridicule their students. | 2.79 | 1.004 |
| 9 | The teachers are authoritarian (strict and controlling) | 2.51 | 1.023 |
| 18 | The teachers appear to have effective communication skills with students | 2.74 | 0.915 |
| 29 | The teachers are good at providing feedback to students | 2.37 | 0.961 |
| 32 | The teachers provide constructive criticism | 2.59 | 0.895 |
| 37 | The teachers give clear examples | 2.96 | 0.758 |
| 39 | The teachers get angry in teaching sessions. | 2.44 | 1.043 |
| 40 | The teachers are well prepared for their teaching sessions | 3.01 | 0.874 |
| 50 | The students irritate the teachers. | 2.33 | 1.040 |
| **Students' academic self-perception (SASP)** |  |  |  |
| 5 | Learning strategies which worked for me before continue to work for me now | 2.10 | 1.207 |
| 10 | I am confident about passing this year. | 3.18 | 0.883 |
| 21 | I feel I am being well prepared for my profession | 2.59 | 0.992 |
| 26 | Last year's work has been a good preparation for this year's work | 2.52 | 0.996 |
| 27 | I am able to memorize all I need. | 2.02 | 1.050 |
| 31 | I have learned a lot about empathy in my profession | 2.53 | 0.963 |
| 41 | My problem solving skills are being well developed here. | 2.70 | 0.903 |
| 45 | Much of what I have to learn seems relevant to a career in healthcare. | 3.03 | 0.876 |
| **Students' perception of the atmosphere (SPA)** |  |  |  |
| 11 | The atmosphere is relaxed during teaching. | 2.51 | 0.882 |
| 12 | The course is well timetabled | 2.56 | 1.063 |
| 17 | Cheating is a problem in my courses | 2.36 | 1.390 |
| 23 | The atmosphere is relaxed during lectures/teaching | 2.61 | 0.857 |
| 30 | There are opportunities for me to develop interpersonal skills | 2.81 | 0.818 |
| 33 | I feel comfortable in teaching sessions socially. | 2.56 | 0.916 |
| 34 | The atmosphere is relaxed during tutorials | 2.72 | 0.767 |
| 35 | I find experience disappointing | 2.55 | 0.850 |
| 36 | I am able to concentrate well | 2.38 | 0.995 |
| 42 | The enjoyment outweighs the stress of studying medicine | 1.39 | 1.104 |
| 43 | The atmosphere motivates me as a learner | 2.22 | 1.047 |
| 49 | I feel able to ask the questions I want | 2.48 | 1.044 |
| **Students' social self-perception (SSSP)** |  |  |  |
| 3 | There is a good support system for students who get stressed. | 2.02 | 1.050 |
| 4 | I am too tired to enjoy this course. | 2.32 | 0.997 |
| 14 | I am rarely bored on this course | 1.81 | 1.113 |
| 15 | I have good friends in this course | 3.01 | 0.892 |
| 19 | My social life is good. | 2.41 | 0.948 |
| 28 | I seldom feel lonely. | 2.25 | 1.129 |
| 46 | My accommodation is pleasant. | 2.05 | 1.186 |

Across the five DREEM domains, the mean scores were as follows (Table 4):

**Table 4.** Summary of DREEM domains.

| **Domains** | **SPL** | **SPT** | **SASP** | **SPA** | **SSSP** | **DREEM** |
| --- | --- | --- | --- | --- | --- | --- |
| Mean | 29.0 | 30.0 | 20.7 | 29.1 | 15.9 | 124.6 |
| Std. Deviation | 7.72 | 5.90 | 5.67 | 6.51 | 4.01 | 23.65 |

Students’ Perception of Learning (SPL): 29.0 ± 7.72

Students’ Perception of Teachers (SPT): 30.0 ± 5.90

Students’ Academic Self-Perception (SASP): 20.7 ± 5.67

Students’ Perception of Atmosphere (SPA): 29.1 ± 6.51

Students’ Social Self-Perception (SSSP): 15.9 ± 4.01

The overall DREEM score was 124.6 ± 23.65, which indicates a “more positive than negative” perception of the learning environment according to standard DREEM interpretation (101–150 range).

### Associations between sociodemographic and DREEM domains (Table 5)

Age: Significant differences were found in academic self-perception (p = 0.024), atmosphere (p = 0.036), and social self-perception (p = 0.033), with older students (27–31 years) having higher mean scores. This suggests maturity may positively influence coping and perception of the learning environment.

**Table 5.** Comparison of DREEM domains across sociodemographic and academic variables.

| Independent Variable | Category | SPL<br>Mean<br>(SD) | SPT<br>Mean<br>(SD) | SASP<br>Mean<br>(SD) | SPA<br>Mean<br>(SD) | SSSP<br>Mean<br>(SD) | DREEM<br>Mean<br>(SD) |
| --- | --- | --- | --- | --- | --- | --- | --- |
| Age | 17 - 21 | 28.9<br>(7.22) | 29.8<br>(5.75) | 20.1<br>(5.41) | 28.0<br>(6.37) | 15.1<br>(4.19) | 121.9<br>(23.02) |
|  | 22 - 26 | 28.9<br>(8.08) | 29.8<br>(6.10) | 20.8<br>(5.80) | 29.8<br>(6.38) | 16.6<br>(3.68) | 126.0<br>(23.82) |
|  | 27 - 31 | 30.7<br>(9.79) | 34.5<br>(2.88) | 26.5<br>(4.37) | 34.7<br>(7.50) | 16.3<br>(5.08) | 142.7<br>(23.77) |
|  | *H (p value)* | 1.428<br>(0.490) | 4.712<br>(0.095) | 7.476<br>(0.024) | 6.667<br>(0.036) | 6.808<br>(0.033) | 5.293<br>(0.071) |
|  |  |  |  | * | * | * |  |
| **Gender** | Male | 29.8<br>(7.31) | 30.4<br>(5.77) | 31.41<br>(5.39) | 30.0<br>(5.90) | 16.3<br>(3.73) | 128.0<br>(21.13) |
|  | Female | 27.1<br>(8.38) | 28.9<br>(6.12) | 18.9<br>(5.98) | 27.0<br>(7.41) | 14.8<br>(4.48) | 116.7<br>(27.4) |
|  | ***U (p value)*** | 2725.5<br>(0.007)<br>* | 3107.0<br>(0.120) | 2730.0<br>(0.007)<br>* | 2797.5<br>(0.013)<br>* | 2980.5<br>(0.053) | 2704.0<br>(0.006)* |
| **Department** | Medicine<br>and Surgery | 29.4<br>(7.11) | 30.3<br>(5.72) | 21.1<br>(5.24) | 29.5<br>(6.16) | 16.0<br>(3.84) | 126.2<br>(22.01) |
|  | Dentistry | 26.8<br>(10.30) | 28.3<br>(6.64) | 18.6<br>(7.36) | 27.4<br>(8.06) | 15.0<br>(4.83) | 116.1<br>(30.12) |
|  | ***U (p value)*** | 2061.0<br>(0.390) | 1877.0<br>(0.121) | 1826.5<br>(0.082) | 1955.0<br>(0.224) | 1989.5<br>(0.259) | 1836.5<br>(0.090) |
| **Academic**<br>**level** | 2nd year | 27.8<br>(8.09) | 29.1<br>(6.29) | 19.7<br>(6.17) | 28.1<br>(6.23) | 15.0<br>(4.09) | 119.7<br>(24.02) |
|  | 3rd year | 30.1<br>(7.19) | 30.8<br>(5.39) | 21.6<br>(4.97) | 30.2<br>(6.65) | 16.8<br>(3.75) | 129.5<br>(22.33) |
| | ***U (p value)*** | 3551.0<br>(0.026)<br>* | 3644.0<br>(0.049)<br>* | 3554.0<br>(0.027)<br>* | 3448.5<br>(0.013)<br>* | 3163.0<br>(0.001)<br>* | 3104.0<br>($<0.001$)<br>* |
| **Accommoda**<br>**tion type** | Off campus | 28.9<br>(7.70) | 29.8<br>(5.63) | 19.1<br>(5.81) | 26.6<br>(6.51) | 15.2<br>(4.37) | 119.6<br>(24.95) |
|  | On campus | 29.0<br>(7.75) | 30.0<br>(5.98) | 21.0<br>(5.60) | 29.7<br>(6.40) | 16.0<br>(3.93) | 125.8<br>(23.28) |
|  | ***U (p value)*** | 2595.5<br>(0.985) | 2581.5<br>(0.945) | 2041.0<br>(0.049) | 1879.0<br>(0.11) | 2252.0<br>(0.220) | 2231.0<br>(0.195) |
| **Mental**<br>**health**<br>**condition** | No | 29.4<br>(7.24) | 30.1<br>(5.66) | 20.9<br>(5.47) | 29.4<br>(6.32) | 16.0<br>(3.90) | 125.7<br>(22.51) |
|  | Yes | 22.8<br>(12.16) | 28.5<br>(9.20) | 17.6<br>(7.89) | 25.3<br>(8.51) | 13.5<br>(5.15) | 107.6<br>(34.49) |
|  | ***U (p value)*** | 616.5<br>(0.043) | 866.0<br>(0.557) | 758.0<br>(0.226) | 694.0<br>(0.115) | 591.0<br>(0.030) | 675.5<br>(0.093) |
|  |  | * |  |  |  | * |  |
| **Academic grade** | < 50 % | 11.0<br>(4.24) | 17.5<br>(10.61) | 8.0<br>(8.49) | 14.0<br>(4.24) | 10.0<br>(8.48) | 60.50<br>(10.61) |
|  | 50 - 59 % | 28.3<br>(7.47) | 28.6<br>(5.41) | 19.0<br>(5.45) | 27.1<br>(6.41) | 15.9<br>(3.82) | 118.9<br>(23.65) |
|  | 60 - 69 % | 27.9<br>(8.29) | 30.8<br>(5.90) | 19.5<br>(6.31) | 28.6<br>(6.61) | 15.3<br>(4.81) | 122.1<br>(25.78) |
|  | ≥ 70 | 30.0<br>(7.14) | 30.2<br>(5.73) | 21.8<br>(4.93) | 30.2<br>(6.08) | 16.2<br>(3.53) | 128.4<br>(20.75) |
|  | **H (p value)** | 7.878<br>(0.049)<br>* | 7.602<br>(0.055) | 14.034<br>(0.003)<br>* | 10.191<br>(0.017)<br>* | 2.221<br>(0.528) | 9.586<br>(0.022)* |
| **Kessler scale** | well | 29.5<br>(7.43) | 30.8<br>(6.20) | 21.3<br>(5.85) | 30.1<br>(6.23) | 16.5<br>(3.79) | 128.2<br>(24.02) |
|  | mild distress | 28.5<br>(8.00) | 30.0<br>(5.22) | 20.6<br>(5.36) | 28.7<br>(6.33) | 15.5<br>(4.07) | 123.3<br>(22.44) |
|  | moderate distress | 32.4<br>(5.14) | 31.0<br>(5.09) | 20.7<br>(5.14) | 31.0<br>(5.31) | 16.4<br>(3.00) | 131.6<br>(16.42) |
|  | severe distress | 25.1 | 26.3 | 18.9 | 25.2 | 14.1 | 109.7 |
|  | **H (p value)** | 10.595<br>(0.014)<br>* | 15.069<br>(0.002)<br>* | 3.526<br>(0.317) | 13.262<br>(0.004)<br>* | 7.924<br>(0.048)<br>* | 14.601<br>(0.002)* |

U = Mann Whitney U score; H = Kruskal Wallis score; \* = p value is significant

Gender: Male students reported significantly higher scores in SPL (p = 0.007), SASP (p = 0.007), SPA (p = 0.013), and overall DREEM (p = 0.006). This may reflect gender-based differences in adaptation, confidence or perceived inclusivity in the learning environment.

Department: No statistically significant differences were found between Medicine and Dentistry students, though medical students tended to have slightly higher mean scores across all domains.

Academic Level: Third-year students reported significantly higher scores across all domains (p < 0.05). This suggests that familiarity with the learning system and increased clinical exposure might enhance perception of teaching, atmosphere, and self-confidence.

Accommodation Type: No significant difference was observed in most domains, though on-campus students reported slightly higher mean DREEM scores (125.8 vs. 119.6), possibly reflecting easier access to academic and social support.

Mental Health Condition: Students without a mental illness scored significantly higher in SPL (p = 0.043) and SSSP (p = 0.030). This indicates that psychological wellbeing positively affects engagement and social integration in the learning environment.

Academic Grade: Students with higher grades (≥70%) reported significantly better SPL (p = 0.049), SASP (p = 0.003), SPA (p = 0.017), and overall DREEM (p = 0.022) scores, suggesting a positive link between academic performance and perception of the learning environment.

Psychological Distress (Kessler Scale): Students classified as *well* or *mildly distressed* had significantly higher scores across SPL (p = 0.014), SPT (p = 0.002), SPA (p = 0.004), SSSP (p = 0.048), and DREEM (p = 0.002) compared to those with *severe distress*. This shows that higher psychological distress is associated with poorer perception of the learning environment.

### Correlation between psychological distress and DREEM domains (Table 6)

There was a significant negative correlation between students’ psychological distress and their overall perception of the learning environment (r = –0.162, *p* = 0.027). Also, psychological distress was significantly associated with poorer perception of teachers (r = – 0.174, p = 0.017), atmosphere (r = –0.158, p = 0.030) and social self-perception (r = –0.177, p = 0.015). Although the correlations with academic self-perception (r = –0.143, p = 0.051) and perception of learning (r = –0.057, p = 0.438) were not statistically significant, the negative direction of all coefficients indicates a consistent trend of psychological distress adversely influencing how students perceive various aspects of their educational environment.

**Table 6.** Correlation between psychological distress and DREEM domain scores.

| DREEM domains | SPL_total | SPT_total | SASP_total | SPA_total | SSSP_total | DREEM |
| --- | --- | --- | --- | --- | --- | --- |
| Correlation coefficient | -0.057 | -0.174 | -0.143 | -0.158 | -0.177 | -0.162 |
| P value | 0.438 | 0.017 | 0.051 | 0.030 | 0.015 | 0.027 |

## Discussion

This study evaluated medical and dental students’ perception of their learning environment using the DREEM inventory and explored the relationship between these perception, sociodemographic characteristics, and psychological distress. The overall DREEM score indicated that students generally viewed their learning environment as more positive than negative. However, findings from the Kessler Psychological Distress Scale revealed that over half of the respondents experienced some degree of psychological distress, with an average score suggestive of moderate stress levels. Moreover, psychological distress demonstrated a significant negative correlation with the total DREEM score and several of its domains, underscoring the potential influence of mental wellbeing on how students perceive and engage with their educational environment.

Our study revealed a notable level of psychological distress among medical students, suggesting that emotional strain is a common experience within the learning environment. This burden appears higher than what has been reported among medical students in Japan (32), Pakistan (33), and India (34), indicating that contextual factors such as resource limitations, curricular pressure, and inadequate mental health support may play a significant role. The finding reinforces the growing recognition that the rigours of medical education in low- and middle-income countries can have profound psychological implications, emphasising the need for early identification and institutional strategies to foster student wellbeing.

The overall DREEM score in this study indicates that students held a generally positive view of their learning environment. This suggests that, despite the challenges inherent in medical education, the preclinical school learning environment is perceived as moderately conducive to learning. Compared with earlier research among medical students in Nigeria (24,25,35–37), our result is higher, although most of those studies were not conducted specifically within preclinical settings. Similarly, it surpasses findings reported in several African countries such as Uganda (38) and Ghana (39), as well as in other contexts including Spain (40), Pakistan (41), Hungary (42), Morocco (43), and Chile (44). However, the score remains lower than those observed in Malaysia (39,45), Thailand (46), Sweden (47), and by Idon *et al*. in Nigeria (48). These cross-country variations may reflect differences in curriculum design, teaching methods, institutional culture, and available academic support systems, all of which influence how students perceive and engage with their learning environment.

### Students’ perception of learning

The SPL domain score reflects a generally positive approach to learning, consistent with reports from other Nigerian medical schools (24,25,36). Interestingly, students perceived the teaching methods as simultaneously student-centred and teacher-centred, suggesting the presence of a hybrid instructional model. While this indicates that the teaching strategies employed foster competence, confidence, and learner satisfaction, it also reveals that certain aspects of the learning experience remain predominantly teacher-centred and may require review. This aligns with the findings of Lateef et al., who similarly reported that medical education is perceived as both student- and teacher-centred (49), reflecting a transitional phase in pedagogical practice within the Nigerian medical education context.

### Students’ perception of teachers

The students’ perception of teachers indicates a generally positive trend. Teachers were viewed as knowledgeable, well prepared for teaching sessions, and able to illustrate concepts with clear examples. These qualities reflect a commendable level of professional competence and commitment to effective instruction which have been reported in other Nigerian medical schools (25,36), suggesting that medical educators across different settings are generally regarded as content experts who deliver teaching effectively. However, some areas warrant attention. The perception of teachers as authoritarian and occasionally ridiculing students suggests relational barriers that could hinder open communication and psychological safety within the learning environment. Furthermore, feedback and constructive criticism were identified as weaker aspects, highlighting the need for enhanced mentorship skills and more supportive teacher–student interactions.

### Students’ academic self-perception

The students’ academic self-perception was generally positive. They expressed confidence in their ability to succeed academically and considered the curriculum relevant to their future professional roles. The learning environment was also perceived as supportive of the development of problem-solving skills, reflecting an encouraging educational atmosphere. However, challenges were noted in adapting learning strategies, as previously effective approaches appeared less suitable for current academic demands. Difficulties with memorization were also highlighted, suggesting that the volume and complexity of material may overwhelm conventional study habits. There is therefore a need for deliberate efforts to help students cultivate adaptive and dynamic learning strategies as they progress through different stages of medical education, recognising that learning contexts and cognitive demands evolve over time.

### Students’ perception of the atmosphere

The preclinical school is generally perceived as a positive atmosphere. The students’ perception indicates that the general classroom environment is conducive to learning. Students reported feeling relatively comfortable and able to engage during teaching sessions, suggesting that interpersonal dynamics within the classroom are largely supportive. Nonetheless, certain aspects detract from this favourable perception. Persistent academic stress appears to overshadow enjoyment, and reduced concentration and motivation may reflect the demanding and high-stakes nature of medical training. While opportunities for interpersonal skill development exist, the overall atmosphere may still benefit from greater attention to student wellbeing, workload management, and interactive learning experiences that sustain motivation and reduce burnout risk.

### Students’ social self-perception

The students’ social self-perception reveals notable gaps in the supportive and social dimensions of the learning environment. While friendships among peers appear strong and serve as informal sources of encouragement, there is a clear absence of an effective institutional support system for students who experience stress. This lack of structured psychosocial assistance suggests that students may be left to rely largely on peer relationships rather than organised mechanisms of support such as counselling or mentorship programmes (18). Furthermore, the frequent feelings of boredom reported among students indicate that learning experiences may not always be stimulating or varied enough to sustain engagement. The perception of poor accommodation further compounds these challenges, as an uncomfortable living environment can heighten fatigue and stress, reducing students’ ability to fully participate and thrive academically. Together, these factors portray a social environment that, while rich in peer connection, falls short in providing the institutional and infrastructural support necessary for optimal student wellbeing.

### DREEM inventory, academic characteristics, sociodemographic characteristics and psychological distress

The findings from this study revealed that increasing age (27–31 years) was significantly associated with higher academic self-perception (p = 0.024), possibly due to better coping strategies among older students. However, a 2019 study reported that younger medical students tend to perform better in final preclinical examinations than their older counterparts, suggesting that the present finding may reflect confidence and self-belief rather than actual academic performance (50). In Nigeria, it is common for secondary school leavers to struggle for some years to gain admission into competitive courses such as Medicine, Surgery, and Dentistry which may make the older entrants have greater positive self-perception (51). A similar trend was observed with perception of atmosphere (p = 0.036), as students who have waited longer to gain admission may feel more appreciative of their environment and less likely to complain about issues such as infrastructure or teaching conditions. Social self-perception was also higher among older students (p = 0.033), which aligns with the general observation that older individuals tend to find it easier to relate with others and build social connections.

These patterns carry practical meaning as students who arrive after a long wait often bring resilience, clearer goals and a strong motivation to make the most of scarce opportunities. Such attributes can colour how they perceive teaching and the learning climate, so their higher scores may reflect gratitude and determination as much as measured teaching quality. Recognising this helps interpret DREEM scores more wisely as a high academic self-perception among this demographic is not automatically proof of superior pedagogy, but a valuable indicator of student mindset and potential engagement.

Some previous studies report higher DREEM scores for females, others for males, and overall there is no clear consensus (24,52,53). Interestingly, males in this study had higher overall DREEM scores and higher scores across the different subscales, which was also statistically significant (DREEM p = 0.006). This could be explained by the tendency of males to adapt more quickly to new situations compared to females (54). It may also relate to the fact that clinical training is more hands-on, and many female students often express a stronger preference for clinical training where they tend to show more empathy, which can improve their perception in the clinical years compared with the preclinical phase where theoretical reading and long hours of study dominate (55). This difference may therefore reflect the nature of preclinical learning rather than any inherent gender differences.

It is also not surprising that third-year students recorded higher DREEM scores (p < 0.05). Having been in the system for a longer period, they would have found their footing academically, socialised more with peers and teachers, and become more accustomed to the atmosphere. The familiarity that comes with time often fosters a sense of belonging and confidence, which positively influences perception of the learning environment. This underlines the value of early orientation and structured social integration activities for first- and second-year students so they can reach that same comfort level sooner.

Although not statistically significant, students staying off-campus recorded lower scores. This may be attributed to the geographical spread of students residing off-campus, which makes it less convenient for them to form tutorial groups, friendship networks, or social support systems. In contrast, those living within the institutional facilities have easier access to colleagues, can meet more frequently, and are more likely to engage in collaborative learning and social activities that enrich their academic experience.

Mental health conditions are significant stressors that affect all aspects of wellbeing (56,57). It is not unexpected that those with mental health challenges had lower self-perception of learning (SPL p = 0.043). The stress associated with illness management, alongside the demands of medical training, can make concentration difficult. Attending classes while experiencing even mild symptoms can significantly impair learning effectiveness and academic engagement. This is an urgent signal for medical schools that mental health support is not optional. In addition, students with higher grades had higher SPL and overall DREEM scores (DREEM p = 0.022), which is understandable since satisfactory academic performance often compensates for perceived weaknesses in the learning environment.

Finally, the findings from the Kessler Distress Scale reinforce the intricate link between mental health and perception of the learning environment. A 2022 study among Malaysian medical students reported an inverse relationship between DREEM scores and likelihood of depression (58). Severe psychological distress negatively affects students’ perception of learning, atmosphere, academic performance and social self-perception (DREEM p = 0.002). This is consistent with the results of this study, which revealed that students experiencing higher levels of distress reported poorer overall perception as well as negative correlation between psychological distress and perception of the learning environment.

Psychological wellbeing therefore plays a crucial role in shaping how students experience and evaluate their educational environment, highlighting the need for adequate mental health support within medical training programmes. In short, the results are not just descriptive; they point to actionable priorities. By translating these insights into policy and practice, institutions can create a learning environment that is not only perceived as better but truly functions better for every student.

### Strengths and limitations

This study offers several noteworthy strengths. First, it focuses specifically on preclinical medical students, a group often overlooked in studies assessing the learning environment, even though this stage forms the foundation of students’ professional and academic development.

Second, it is the first known study in the Nigerian context to examine the perception of the learning environment using the DREEM tool in relation to psychological distress, providing novel insight into the intersection between academic conditions and mental wellbeing. In addition, the use of validated instruments enhances the reliability and comparability of the findings with international research.

Despite its strengths, this study has some limitations. It was conducted in a single medical school, which may limit the generalisability of the findings to other institutions with different curricula or learning contexts. Also, the study relied on self-reported data, which may be influenced by social desirability or recall bias. Additionally, contextual factors such as examination schedules, institutional policies, or external stressors during the study period may have influenced students’ perception at the time of data collection. However, the analytic design compensated for the cross-sectional study design limitation by establishing strong associations between psychological well-being of students and the learning milieu.

### Recommendations

To enhance the learning environment of medical and dental students, institutions should prioritise structured academic and social integration from the earliest stages of training. Mentorship schemes that link junior students with senior colleagues can help build confidence and maturity. Early orientation programmes, guided campus tours, and social mixers with faculties and senior students will also help new entrants feel grounded and connected. Regular, low-stakes opportunities for engagement with staff and peers, such as informal discussions, peer-led tutorials, and study groups, can promote a sense of belonging and reduce early-year feelings of uncertainties and mixed emotions. Learning methods should be diversified beyond lectures. Increasing preclinical practical exposure and using varied assessment formats will support students with different learning preferences. Institutions should also aim to accommodate all medical and dental students on campus, as being close to academic and social facilities encourages participation and collaboration.

Mental well-being should be treated as a core part of the educational experience. Early screening and mental health awareness should be embedded in the curriculum. Staff should be trained to recognise and respond to signs of distress. Structured study-skills workshops, formative assessments with constructive feedback, and mentoring schemes should be provided regularly to build competence, confidence, and resilience. Future research should explore how students’ perception change as they move into clinical years and examine the impact of targeted interventions on their long-term academic and professional outcomes.

## Conclusion

Preclinical medical and dental students at the University of Ibadan perceive their learning environment as more positive than negative. However, a significant number experience considerable psychological distress, which calls for urgent intervention by medical educators, institutions, and policymakers. These experiences can shape students’ attitudes as they progress into clinical training and ultimately influence their professional competence and career fulfilment. Also, helping students build strong social and emotional intelligence during the preclinical years can enhance resilience, interpersonal relationships, and adaptability, thereby strengthening academic outcomes. Evidence-based reforms are needed to foster a healthier, more inclusive, and productive learning environment for future health professionals.

## DECLARATIONS ACKNOWLEDGEMENT

Nil

## ETHICS APPROVAL AND INFORMED CONSENT TO PARTICIPATE

The study received approval from the University of Ibadan / University College Hospital (UI/UCH) institutional review board with IRB number 25/0418. Informed consent to participate was obtained from all participants prior to their inclusion in the study in accordance with ethical guidelines and the Declaration of Helsinki.

## CONSENT FOR PUBLICATION

All participants provided consent for the anonymous use of their data for research dissemination and publication purposes. No identifiable personal information was collected or reported.

## AVAILABILITY OF DATA AND MATERIALS

This will be made available on request from the corresponding author.

## CLINICAL TRIAL NUMBER

Not Applicable

## CONTRIBUTORS

**AAA** conceived the study, designed the research protocol, performed the data analysis, and wrote the results and discussion sections and participated in data collection.

**AOO** contributed to the study conception and design, wrote the introduction and part of the discussion, and participated in data collection.

**MOO** participated in data collection and wrote the methods section.

**MD** participated in data collection and co-wrote the methods section.

**EOI** provided overall supervision throughout the study period, critically reviewed the manuscript, and approved the final version.

All authors read and approved the final manuscript.

## Data Availability

All data produced in the present study are available upon reasonable request to the authors

